# Three decades of hospital evacuations in the Netherlands: a scoping review

**DOI:** 10.1101/2022.01.31.22270213

**Authors:** Dennis G. Barten, Marjolein H.M. Fijten, Menno I. Gaakeer, Vincent W. Klokman, Luc J. Mortelmans, Frits van Osch, Nathalie A.L.R. Peters, Jaap J.J. Wijnands, Edward C.T.H. Tan, Arjen Boin

**Author notes:** **Corresponding author:** D.G. Barten, MD, Department of Emergency Medicine, VieCuri Medical Center, P.O. Box 1926, 5900 BX Venlo, The Netherlands.

## Abstract

**Background:** Hospitals are expected to provide their essential services under all circumstances, including disasters and crisis situations. However, crises and disasters may disrupt essential hospital functions, and necessitate evacuation of the facility. Data on the prevalence and causes of hospital evacuations is limited, whilst realistic evacuation planning will make hospitals more resilient during future crises and disasters. The aim of this study was to develop a systematic understanding of hospital evacuation frequency, causes and characteristics in the Netherlands.

**Methods:** A systematic scoping review of news articles retrieved from the LexisNexis database, Google, Google News, PubMed and EMBASE between 1990 and 2020. All articles mentioning a hospital evacuation in the Netherlands were analyzed.

**Results:** A total of 67 hospital evacuations were identified in a 30-year time period. The most common primary incidents were internal fire (33%), technological failure (21%) and hazardous materials (12%). Cascading events transpired in 37% of all events. As a result, internal fire (34%), internal power failure (19%) and hazardous materials (16%) were the most important reasons to evacuate a medical facility. The number of evacuees ranged from 1 to 339 patients (median 10.5, interquartile range 36), and there were 13 (19%) complete hospital evacuations. The frequency of hospital evacuations increased over time, with a more diverse etiology during later years.

**Conclusions:** Hospital evacuations do occur, and hospitals are vulnerable to both internal and external hazards. Internal fire, internal power failure, hazardous materials and flooding incidents were the most important reasons to (partly) evacuate a hospital. Our findings suggest that facility-specific evacuation guidelines are necessary, and that evacuation preparedness will benefit if the most relevant evacuation scenarios are planned and practiced.

## Introduction

Hospitals are expected to provide their essential services under all circumstances, including disasters and crisis situations. However, these very situations may also disrupt hospital functions and subsequently create a threat to patient safety and the continuity of emergency and non-emergency care.^1 2^ Sudden onset events that severely disrupt the everyday, routine services of a hospital facility are referred to as ‘internal hospital crises and disasters’ (IHCDs).^1 3^ IHCD frequency is increasing over time, which is in part explained by the growing impact of information and communication technology (ICT) failures as well as the increasing complexity of hospital infrastructures.^3 4^ A previous study from the Netherlands revealed that the annual risk for a hospital to experience an IHCD was 16% between 2010 and 2020.^3^

There is limited data on the prevalence and causes of hospital evacuations. Vulnerabilities of medical facilities may differ between countries due to certain dominant event types and/or geographical conditions. Whilst in some countries more than 50% of healthcare facilities are situated in high-risk areas for natural disasters, other regions may be more susceptible to man-made disasters such as (cyber)terrorism.^5-7^ The Sendai Framework for Disaster Risk Reduction 2015-2030 encourages to substantially improve resilience to damage to critical infrastructure and disruption of basic services by 2030.^8^ This particularly concerns healthcare and educational facilities. Although a hospital evacuation cannot be avoided at all times, planning and training for hospital evacuation will make it more resilient in the face of future crises and disasters.^5^ IHCDs commonly occur and are frequently associated with the evacuation of patients.^3^ Furthermore, it has been shown that evacuation guidelines could be improved.^5 9^

Learning from past events may help to improve evacuation preparedness. Unfortunately, hospital evacuations are scarcely reported in medical literature, but newspapers, press releases and other gray literature often do include valuable information on these incidents. Therefore, a scoping review strategy was chosen to acquire all available reports of hospital evacuations in the Netherlands from 1990 until 2020, by screening news media, medical literature and gray literature. The objective of this study was to assess the frequency, causes and characteristics of hospital evacuations, as well as to identify temporal trends.

## Materials and methods

A scoping review was conducted following the framework described by Arksey & O’Malley^10^ and reported per PRISMA-ScR guidelines.^11^ This study was a secondary analysis (2000-2020) and extension (1990-2000) of the study by Klokman et al.^3^, who mapped the occurrence of IHCDs in the Netherlands from 2000 until 2020 using the same scoping review strategy. The scoping review protocol was developed *a priori* to guarantee reproducibility and transparency. The protocol, incident definitions, data characterization form and dataset are available upon request. The incident review team consisted of individuals with multidisciplinary expertise in emergency medicine, disaster medicine, public health, methodology, crisis analysis and crisis management. The institutional review board of VieCuri Medical Center approved this research project (#2020_007).

### Search

Hospital evacuations between January 1, 2000 and December 31, 2019 were extracted from the IHCD database of Klokman et al.^3^. For hospital evacuations in the Netherlands between January 1, 1990 and December 31, 1999, an additional search was performed. Public search engines (PubMed, Medline, Google, Google News) and the LexisNexis newspaper article database were used to search for articles and press releases. The search was conducted in June until September of 2020. The search terms “hospital,” “disaster,”, “evacuation”, “moved”, “failure,”, “malfunction”, “fire,”, “smoke”, “explosion”, “flood”, “earthquake”, “bomb threat”, “biologic”, “radioactive”, “nuclear”, “chemical”, “attack” and “nuisance” and their synonyms were combined with Boolean operators.

### Article selection

Articles were only included in this study if the evacuation of one or more hospital wards was mentioned in the article. An evacuation was defined as the process of moving people from one hospital (department) to another hospital (department). Evacuations could occur internally (within the same facility), externally (to another facility) or both. Events were excluded if hospital wards or critical care departments were unaffected or if the event did not take place at an acute care hospital (hospitals containing an emergency department (ED) and intensive care unit (ICU)), if no patients were involved in the evacuation, if the evacuation was planned, and if the evacuation was due to the detection of a multi-resistant micro-organism. The search was conducted by two authors (D.G.B. and M.F.). Duplicates were manually removed. When discrepancies could not be resolved regarding inclusion and characterization, three other reviewers made the final judgement (V.W.K., F.v.O. and A.B.).

### Data collection

Variables that were extracted from the articles included: date and time of onset; date and time of termination; day of the week; season; city and province; impending crisis or disaster (did a disaster actually happen or was there a risk of disaster prompting an evacuation based on the precautionary principle?); internal disaster or combined internal and external disaster; presence of cascading events; primary incident and (when appropriate) final incident; affected hospital departments; number of evacuees and evacuation disposition (internally, externally or both).

### Event classifications

Causes of the hospital evacuation were categorized. In case of cascading events, the events were sequentially noted, with the initial, instigating event defined as the primary incident (PI). The incident that led to the evacuation decision was considered to be the final incident (FI). Each of the events was classified into one of the following categories (also see Appendix 1): technical failure, ICT failure, fire (internal or external), power failure (internal or external), hazardous materials, structural failure, utility failure, loss of medical gases, hydro-meteorological, riverine flooding, flooding due to technical failure, or security & violence.

### Analysis and data charting

All articles mentioning a hospital evacuation were downloaded and data were extracted and independently characterized. Collected data included: location, temporal aspects, failure types, number of evacuees and hospital departments involved. All events were recorded in Castor EDC (Castor Electronic Data Capture, Ciwit BV, Amsterdam, The Netherlands). Statistical analyses were performed using a statistical package program (SPSS version 24, IBM, Chicago, IL.). Descriptive statistics were used to analyse the causes of hospital evacuations over the years. Fisher Exact tests were used to assess possible trends over time. Kruskal-Wallis and Mann-Whitney tests were used to assess possible associations between the number of patients evacuated and other categorical variables. All analyses were conducted with a significance level of P<0.05.

## Results

### Selection of sources of evidence

The scoping review strategy yielded 9.122 records between 1990 and 2000, which resulted in 12 unique hospital evacuation events. The study of Klokman et al. identified 134 IHCDs between 2000 and 2020, of which 55 events involved a hospital evacuation. In total, 67 hospital evacuations between 1990 and 2020 were included in the analysis (Fig 1).

**Fig 1.**
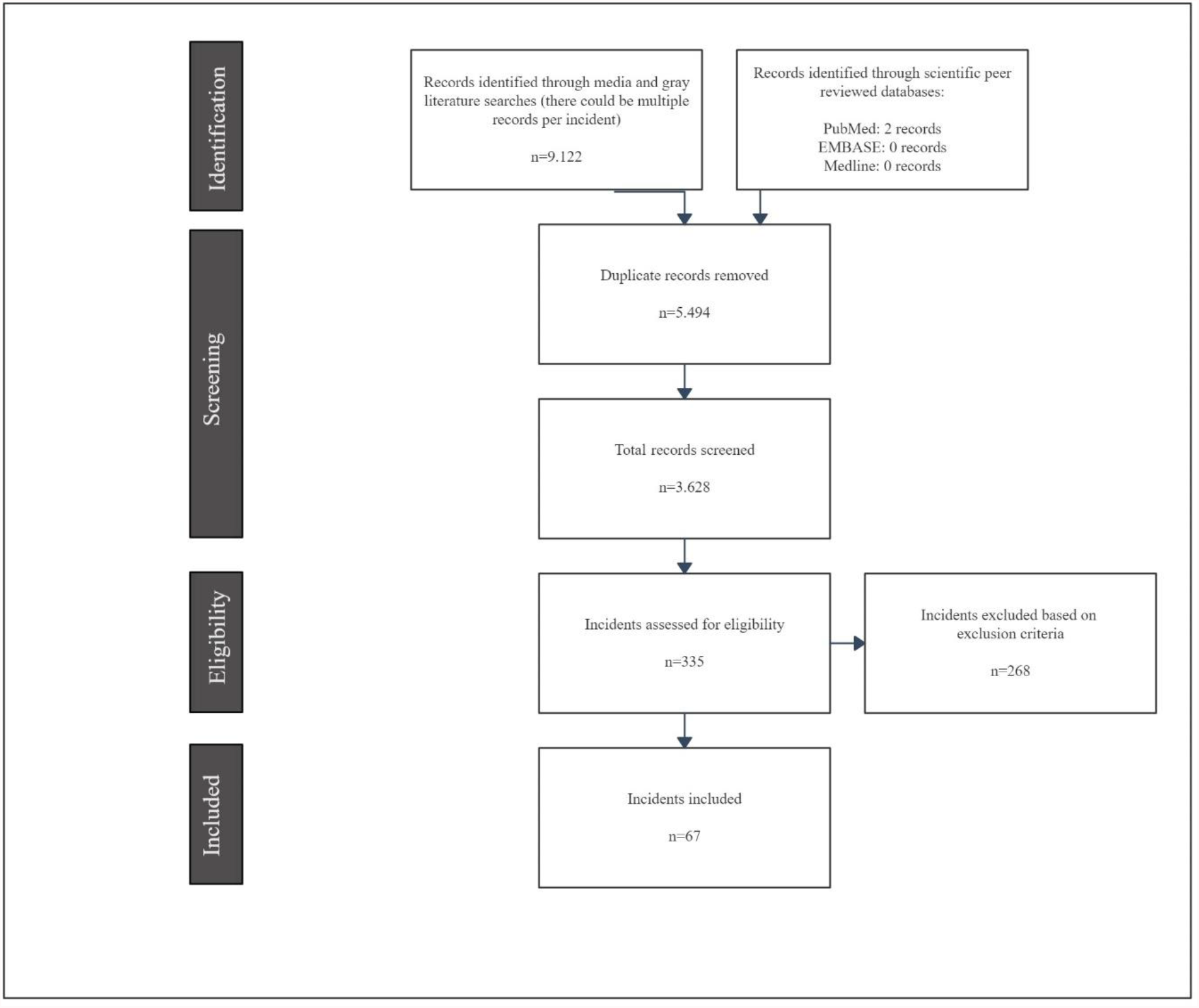
Flow diagram of the scoping review process. Process during the screening and identification of hospital evacuations in the Netherlands between 1990 and 2020.

### Scoping review descriptive statistics

The 67 evacuations in Dutch hospitals occurred in 42 different hospital organisations (mean of 2.2 evacuations per year). All incidents were found in the LexisNexis database and two in PubMed. Twelve evacuations (18%) occurred between 1990 and 2000, 18 evacuations (27%) between 2000 and 2010, and 37 (55%) between 2010 and 2020. This increase over time reached statistical significance (p<0.001). Evacuations were evenly distributed by season and by day of the week. More than half of the evacuations (53%) took place out-of-hours; the onset time was unknown for one event. Ninety-one percent of the evacuations were due to a disaster, whilst the remaining 6 events concerned an impending crisis. Of these, 3 events concerned the risk of imminent riverine floodings. Internal disasters were the most common cause of a hospital evacuation (81%). The remaining events were the result of combined internal and external hospital disasters.

### Primary incident (PI) classification

The most common PIs were internal fire (33%), technological failure (21%), hazardous materials (12%), external power failure (6%), and riverine flooding (6%). The other categories compromised of lower numbers of events (Table 1). Water nuisance incidents (flooding by river, flooding by technical failure and hydro-meteorological) ranked third when combined (13%). The 2 hydro-meteorological incidents concerned flash floods due to extreme precipitation.

**Table 1:** Primary incident classification (n=67)

| Primary incident | Number of events (%) |
| --- | --- |
| Internal fire | 22 (33%) |
| Technological failure | 14 (21%) |
| Hazardous materials | 8 (12%) |
| External power failure | 4 (6%) |
| Flooding by river | 4 (6%) |
| Flooding due to technical failure | 3 (4%) |
| Structural failure | 3 (4%) |
| Internal power failure | 2 (3%) |
| Hydro-metereological | 2 (3%) |
| Security and violence | 2 (3%) |
| External fire | 1 (1%) |
| ICT failure | 1 (1%) |
| Smoke (unknown cause) | 1 (1%) |

The PIs per decade are shown in Fig 2. All riverine flooding events occurred between 1990 and 2000. Half of the evacuations during this first decade were associated with an internal fire as PI, which compares to almost one third of evacuations during the second and third decade. Technical failures were the PI once (8%) during the first decade, but accounted for 44% of events during the second decade and 14% during the third decade. Between 2010 and 2020, hazardous materials were the PI in 16% of evacuations. Furthermore, evacuations with external power failures as PI were only observed during the third decade.

**Fig 2.**
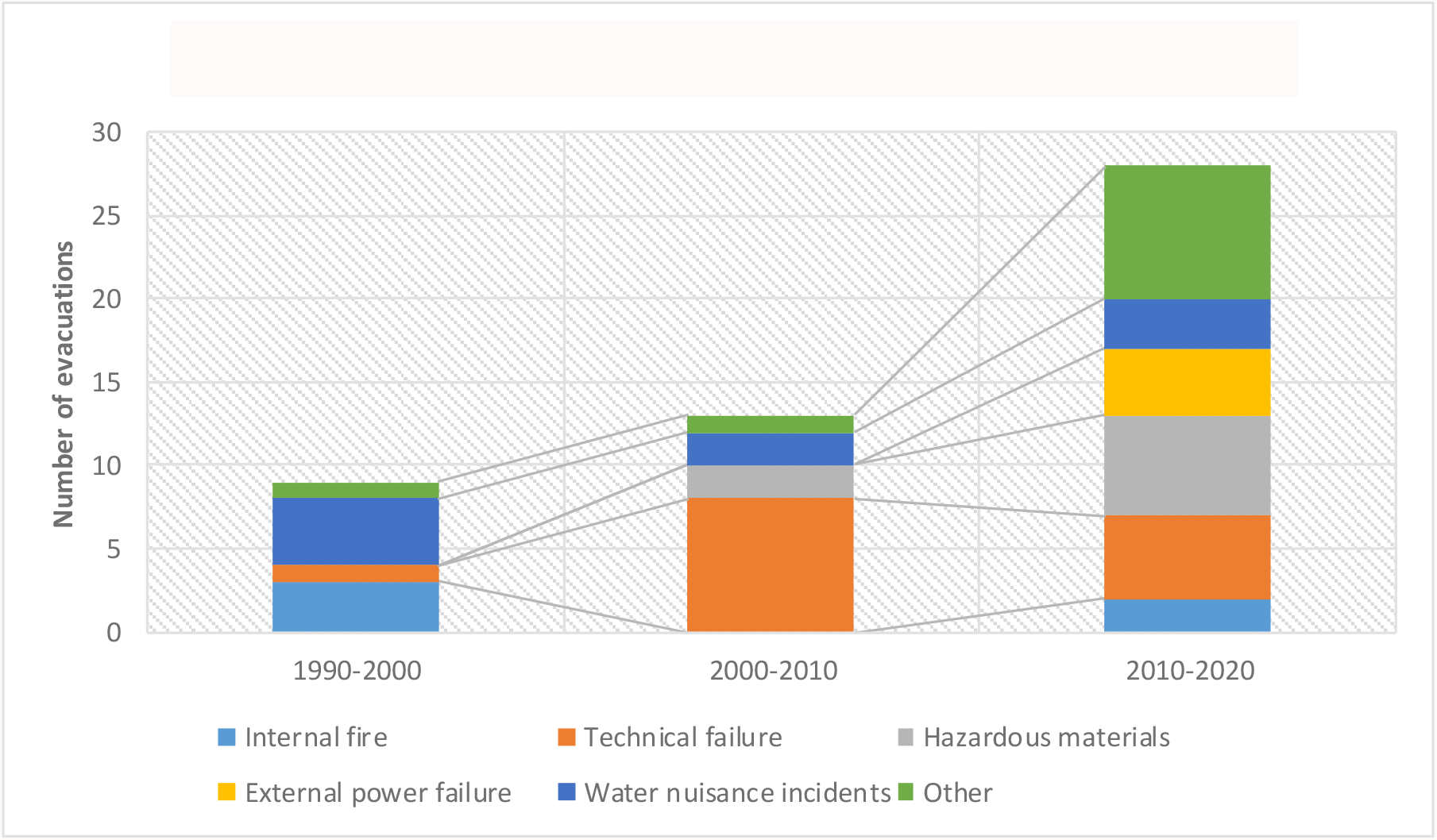
Primary incident classification per decade.

### Cascading events and final incidents

Cascading events were observed in 37% of all events (n=25). Therefore, the PI was not necessarily the final incident (FI) that resulted in evacuation. The most common cascades were internal fire leading to internal power failure (4/25; 16%), external power failure followed by internal power failure (16%) and technical failure resulting in internal fire (16%). All cascading event combinations are presented in Table 2. The cascades related to external power failures are noteworthy: in 3 out of 4 external power failures the emergency generators malfunctioned, necessitating evacuation. When assessing the FI, internal fires (34%) and internal power failures (19%) were most common (Figure 3). Hazardous materials were responsible for 16% and technological failures for 10% of evacuations.

**Table 2:** Cascading events prior to hospital evacuation (n=25)

| Primary incident | Final incident | Number of events (% of total) |
| --- | --- | --- |
| Internal fire | Internal Power Failure | 4 (16.0%) |
| Technical failure | Internal Power Failure | 2 (8.0%) |
| Flooding by river | Internal Power Failure | 1 (4.0%) |
| External power failure | Internal Power Failure | 4 (16.0%) |
| Flooding due to technical failure | Internal Power Failure | 2 (8.0%) |
| Technical failure | Internal Fire | 4 (16.0%) |
| Internal power failure | Internal Fire | 1 (4.0%) |
| Technical failure | Hazardous Materials | 2 (8.0%) |
| Security & violence | Hazardous Materials | 1 (4.0%) |
| Internal power failure | Technical Failure | 1 (4.0%) |
| Hydro-meteorological | Structural Failure | 2 (8.0%) |
| Structural failure | Hydro-meteorological | 1 (4.0%) |

**Figure 3.**
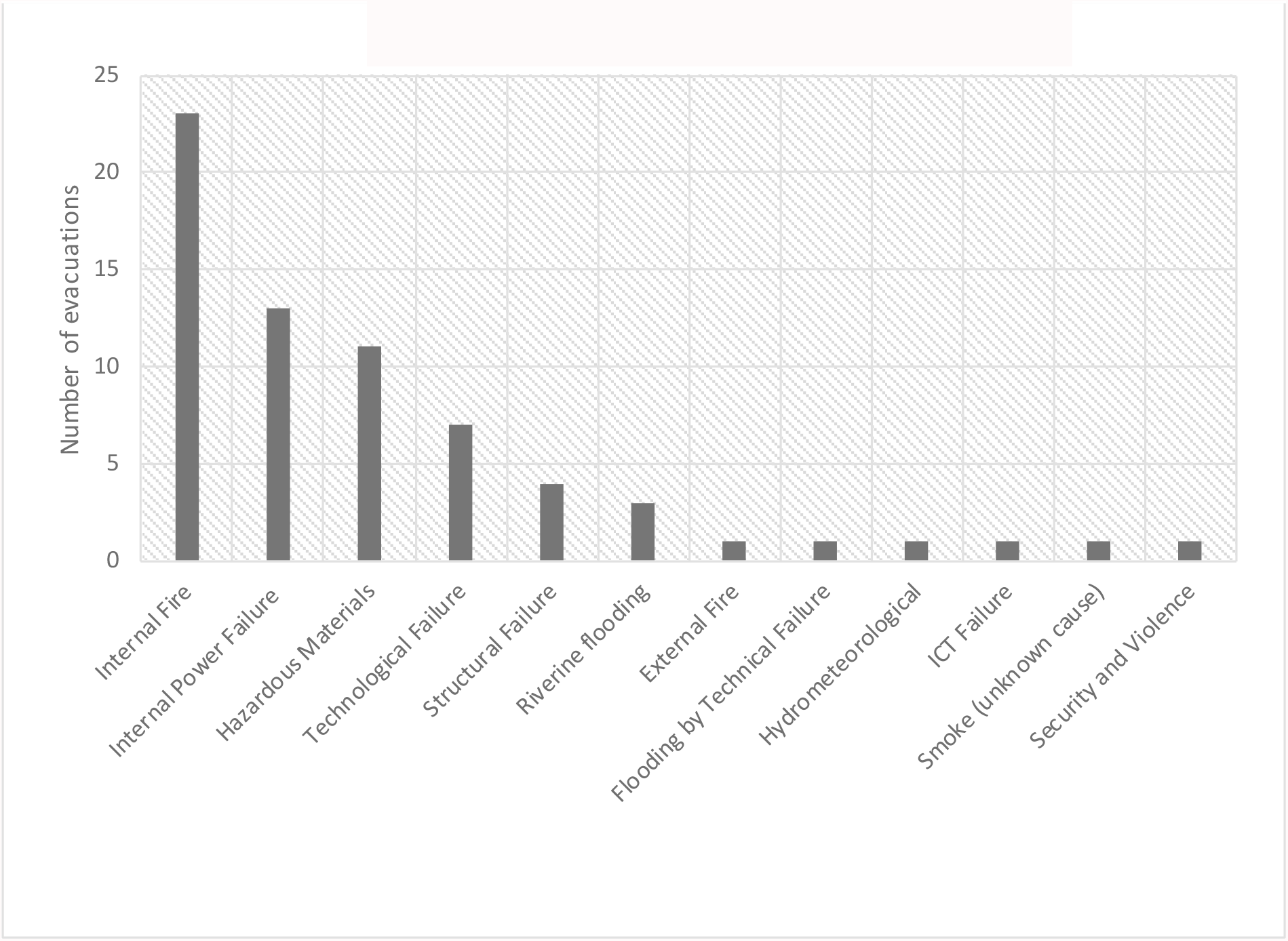
Final incident classification.

### Hospital evacuations: extent and outcomes

On 13 occasions (19%), the entire hospital was evacuated. Evacuations of entire hospitals was associated with the PIs riverine flooding (4/4; 100%), external power failures (3/4; 75%) and flooding by technical failure (2/3; 67%). One hospital was completely evacuated in response to an ICT failure. Multi-departmental evacuations comprised 21% of evacuations and 60% concerned single departments. ED evacuation (40%) was associated with the PIs hazardous materials (6/8; 75%), security & violence (2/2; 100%), internal power failures (2/2; 100%) and hydro-meteorological (2/2; 100%) (p= 0.008).

The number of evacuees was mentioned in 46 events, and ranged from 1 to 339 patients (median 10.5, interquartile range 36). In 7 events, ≥100 patients were evacuated. In 58% of evacuations, patients were evacuated internally, in 25% externally (n= 17). In 16% of the events, patients were both internally and externally evacuated. There was insufficient data to assess the proportion of vertical versus horizontal evacuations.

## Discussion

This study identified 67 hospital evacuations in the Netherlands across a 30-year study period. The most common PIs were internal fire, technological failure and hazardous materials. Cascading events occurred in 37% of all events. As a result, internal fire, internal power failure and hazardous materials were the most important reasons to (partially) evacuate a medical facility. Furthermore, water nuisance incidents (flooding by river, flooding by technical failure and hydro-meteorological combined) were commonly associated with hospital evacuations. The frequency of hospital evacuations increased over time, with a more diverse etiology during later years, likely coinciding with the ever increasing complexity of hospital infrastructure.

Four out of five hospital evacuations in this study resulted from internal hospital disasters. This is in line with a retrospective study of 14 evacuations of medical facilities in Poland, which identified internal fires and bomb threats as the main triggers for evacuation.^12^ Conversely, two other international studies found higher rates of (external) natural disasters, namely earthquakes and hurricanes.^13 14^ One of these studies pertained to the period 1971-1999, largely before the digital revolution healthcare, when the risk of disrupting ICT and power failures was probably lower.^13^ The proportion of water nuisance incidents was significant in this study, but remained steady during the study period. It is very likely that climate change will increase the occurence of future extreme weather events. In the Netherlands, this particularly concerns riverine and flash floods.^1 15^ In 2015, a flood risk analysis determined that 75% of the 185 Dutch hospital locations faced a flooding risk.^6^ Despite these risks, most of the vulnerable hospitals have their ED and emergency generators situated on the ground floor or even in the basement.^6 16^ This suggests that addressing the flood risks of hospitals should be prioritized, especially in flood-prone areas, and that the location of future hospitals should be determined after proper risk analyses of natural and man-made hazards.

Cascading events were frequently observed prior to evacuation decisions, and the PI was not necessarily the FI that resulted in evacuation. Interdependence within complex networks, especially in critical infrastructure networks, increases cascading failure risk and has important implications for infrastructure reliability and security.^17^ Hospitals customarily seem to be able to absorb one “hit” to their internal infrastructure, such as a simple external power failure where the emergency generators seamlessly starts and the hospital continues to function. Conversely, if somewhere in that succession of events an additional failure, such as a technological failure (i.e., short circuit) hindering emergency power supply, a hospital may fail to provide its essential services. Power outages often lead to failures involving computer networks, and technological failures also tend to lead to equipment malfunctions that, in turn, lead to fires and hazardous material releases.^3^ To prevent future IHCDs that necessitate hospital evacuation, one should focus on the prevention of the most common PIs, FIs as well as cascading events. A hospital evacuation for an internal fire carries other challenges than an evacuation procedure for natural disasters, when the external infrastructure and surrounding hospitals may be incapacitated.^1 18^ Furthermore, it is crucial to differentiate between hospital evacuations due to foreseeable threats (i.e. most riverine floods) and those prompted by acute-onset disasters (i.e. earthquakes). This suggests that evacuation planning and practice should take into account various threat scenarios.

The majority of evacuations were completed internally, meaning that hospitals are generally able to accommodate patients within their own facility, especially if the incident is confined to one single department. The sources used in this study lacked detailed information on the disposition of patients, vertical versus horizontal evacuation, and possible challenges. Previous studies highlighted difficulties regarding complete hospital evacuations, particularly with regards to patient disposition, external communication and partnership between hospitals.^5 14^ Furthermore, it was found that there often is insufficient or defective planning for vulnerable groups, including ICU patients.^5^ Triage is another challenge. The typical planning assumption is that all patients will be transferred. However, the availability of resources, as well as time, may not allow for the successful evacuation of all patients. The prioritization of patient evacuation using decision frameworks may reduce morbidity and mortality.^19^ There are specific triage methods for evacuation (Healthcare Evacuation Reverse Triage Priorities), which may ease up the process of selection. However, there is still no common guide for evacuation, and many hospitals lack the proper preparedness.^1 5^ If facility-specific evacuation guidelines are designed, these should incorporate the hospital’s location and surrounding infrastructure, building design, transportation options/routes and the proportion of vertical evacuation of patients in the absence of functioning elevators.^20^

### Limitations

There are several limitations to this review. Under ascertainment may have occurred due to the subjective nature of press and news releases; bias towards sensationalistic and newsworthy events may have arisen, leaving out smaller and less impactful incidents. However, it is likely that events in which a critical care or inpatient department had to be closed and evacuated urgently would have made the news. Underreporting of incidents may have occurred in earlier years due to inaccessibility of articles and decreased number of reporting news outlets. Furthermore, only acute care hospitals were included, and the results cannot be applied to other types of healthcare facilities. Potentially valuable information on details of the evacuation procedure, such as vertical versus horizontal evacuation or the facility type patients were transferred to, were often lacking. Nonetheless, the sources and databases used are the best available. This study was geographically limited to the Netherlands; however, the failure types are likely to be applicable to all modern hospital networks, with the exception of natural disasters, which are more common in some other regions of the world.

## Conclusion

Hospitals are increasingly vulnerable to both internal and external hazards. Hospital evacuations do occur, as we have seen in the Netherlands. Internal fire, internal power failure, hazardous materials and water nuisance incidents were the most important reasons to (partly) evacuate a hospital in the Netherlands. Our findings suggest that facility-specific evacuation guidelines are necessary, and that evacuation preparedness will benefit if the most relevant evacuation scenarios are planned and practiced.

## Data Availability

All data produced in the present study are available upon reasonable request to the authors

## Tables and figures

**Appendix 1:**
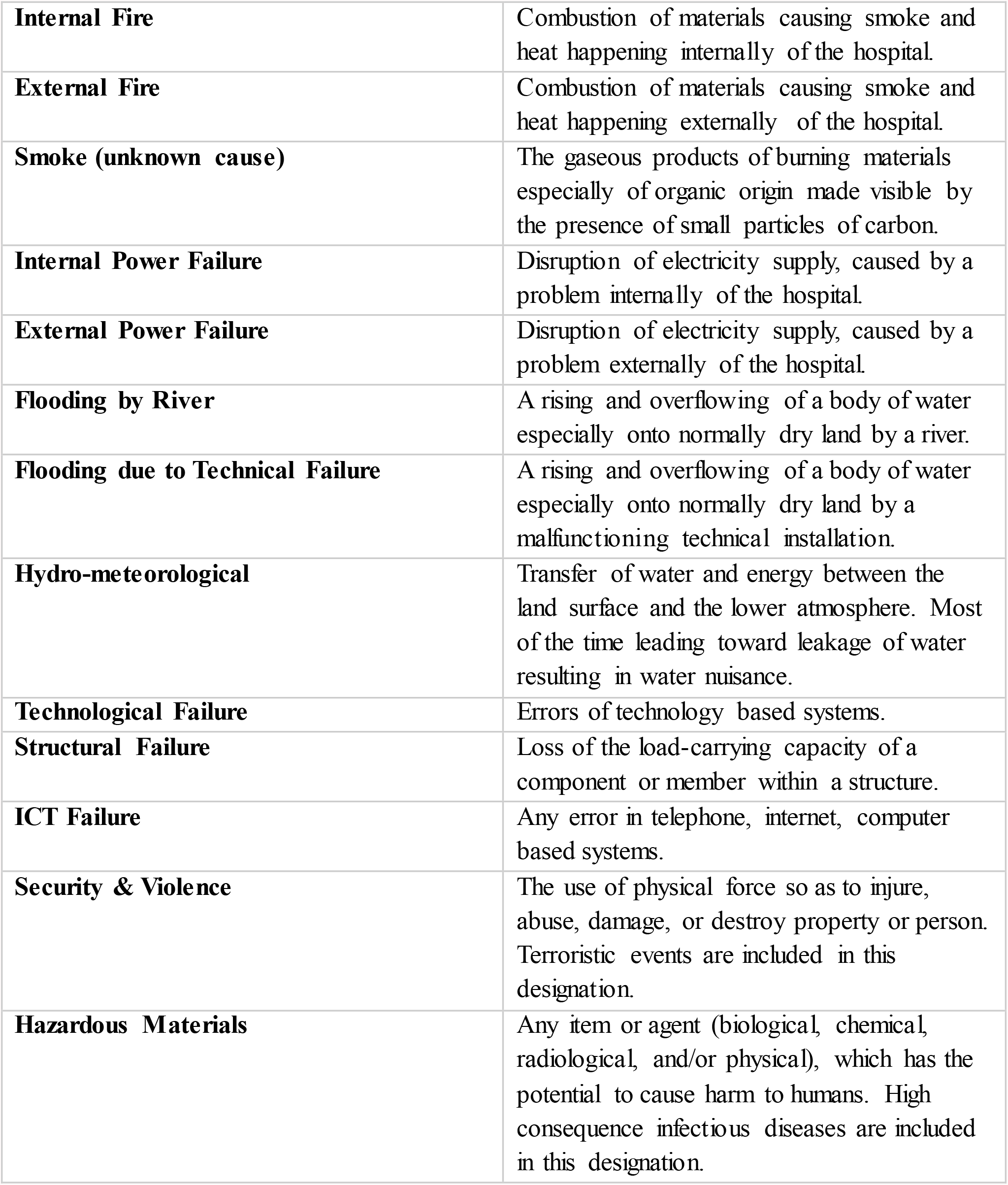
Incident classification.

